# Rational Use of Antibiotics among inpatients at a University Teaching Hospital of Butare, in Rwanda: A Cross-Sectional Study

**DOI:** 10.64898/2026.01.06.26343560

**Authors:** Olivier Icyishatse, Herve Semukunzi, Felix Habarugira, Kato J. Njunwa, Innocent Hahirwa, Jean Baptiste Nyandwi

## Abstract

**Background:** Irrational use of antibiotics is a major factor in antibiotic resistance and poor patient outcomes. Globally, 50% of antibiotics are prescribed inappropriately, in Sub-Saharan Africa alone, antimicrobial resistance was linked to approximately 1.27 million deaths in 2019, mainly due to the misuse and overuse of antibiotics. In this study, we collected data to understand the prescribing patterns and factors associated with irrational antibiotic use among hospitalized patients.

**Methodology:** A hospital-based observational cross-sectional study was conducted between October and December 2024 among 655 patients. The study was conducted at a University Teaching Hospital in Rwanda.

**Results:** Among 655 inpatients, 1,265 antibiotics were prescribed, equivalent to 1.95 per patient. Out of 1463 inpatients in the hospital, 44.76% received at least one antibiotic. The most prescribed antibiotics were Ceftriaxone (34.6%), followed by Flagyl (20%). The majority of antibiotics (83%) were prescribed in injectable forms. Culture and antibiogram testing were performed in 23.20% of cases, this was associated with deviation from national Standard Treatment Guidelines (STGs) adherence (Adjusted OR = 0.07, CI: 0.03-0.14, p = 0.001). there is a significant likelihood of non-adherence to STGs in internal medicine (Adjusted OR=0.25, CI: 0.10-0.62, p = 0.003) and pediatrics (Adjusted OR=0.33, CI: 0.16-0.67, p = 0.002).

**Conclusion:** In this study, we found improper use of antibiotics, deviating from WHO-endorsed standards and a high empiric prescription of antibiotics. Deviation from national STGs was highly associated with empirical prescription. Improving adherence to diagnostic support and antimicrobial stewardship programs could address the problem.

## Introduction

Antibiotics are the most important medicines used in healthcare settings. The discovery of penicillin in 1928 catalyzed the development of numerous antibiotics currently in use for treating different infections [1]. After antibiotics discovery, it was widely believed that infectious diseases would be eradicated. However, the irrational use of these drugs has led to the emergence and spread of antimicrobial resistance [2].

Resistance to antibiotics is estimated to cause approximately 1.27 million deaths worldwide, highlighting its severe global impact [3]. This burden is particularly concerning in developing countries, where antibiotic overuse is widespread, and a significant proportion of prescriptions fail to meet WHO standards for rational use, further accelerating the threat of resistance [4]. The increase of resistance is also involving broad-spectrum antibiotics, including carbapenems [5]. The estimates suggest that almost two million (1.91) deaths would occur due to AMR and 8.22 million deaths of diseases associated with AMR in 2050 [6].

Irrational antibiotic use is commonly manifested as overuse or polypharmacy, non-adherence to guidelines, excessive use of injections, and improper dosing [7]. It was reported that irrational antibiotic use is associated with serious higher risk of adverse effects, prolonged hospital stays, and escalated drug costs [8,9].

According to the WHO, rational antibiotic use involves prescribing the right antibiotic, at the appropriate dose and duration, tailored to the patient’s clinical needs, and at the lowest possible cost to both the patient and the community [10]. However, achieving this standard is often hindered by factors such as limited healthcare provider knowledge, improper procurement, lack of policy and guidelines, and financial constraints [11,12].

Globally, irrational antibiotic use has become a major public health concern. The irrational use of antibiotics is more pronounced in low-and middle-income countries (LMICs) where resources are limited, with over 50% of all antibiotics being inappropriately used [13]. In addition, in these countries, more than 50% of patients are not following their prescribed treatment regimen [14]. The African region alone contributes to approximately 21% of this global irrational antibiotic use [3].

Global responses, including Antimicrobial Stewardship (AMS) programs and the Antimicrobial Resistance Control Program, have shown promising outcomes in promoting rational antibiotic use [15]. WHO recommended strategies such as boosting research and development of novel antibiotics and alternative treatment possibilities, enhancing surveillance and monitoring systems, raising awareness and educating people about the proper antibiotics use and establishing the Essential Medicine Lists (EML) [12].

Despite several measures put in place to enhance the proper use of antibiotics, there is still a surge in irrational use of antimicrobials. A point prevalence survey conducted at Kigali University Teaching Hospital has identified inappropriate prescribing practices, notably the overuse of broad-spectrum antibiotics, with commonly prescribed antibiotics classified in the watch group (65.1%), highlighting significant gaps in antimicrobial stewardship and prescribing protocols [16]. A study reported significant gaps in knowledge and practices concerning antibiotic use in health centers, with evidence pointing to widespread misuse and limited adherence to appropriate prescribing guidelines contributing to antimicrobial resistance [17]

A recent study carried out at the University Teaching Hospital of Butare revealed a concern in resistance of commonly prescribed antibiotics [18]. This pattern indicates a potential misuse or overuse of antimicrobial agents within the clinical setting, which could be contributing to the development of resistant strains.

The rational use of antibiotics in Rwanda, especially at tertiary health care facilities, remains underreported. Thus, this study aimed to evaluate the rational use of antibiotics and identify factors associated with irrational prescribing at the University Teaching Hospital in Rwanda. The findings will contribute to the evidence base needed for strengthening AMS programs and informing national policy. The researcher used data collected prospectively from routine medical records in five major departments of Butare University Teaching Hospital.

## Methods

### Study aim

This study aimed to assess patterns of antibiotic use and identify risk factors contributing to irrational prescribing practices at Butare University Teaching Hospital in Rwanda, intending to inform strategies to tackle antimicrobial resistance

### Study design and setting

An observational cross-sectional study was conducted prospectively between 1 October 2024 and 31 December 2024 among 655 patients. This study was conducted at a University Teaching Hospital in Rwanda, offering specialist medical care, training of medical personnel, thereby contributing to the development of human resource.

### Ethical statement

Ethical approval was obtained from the College of Medicine and Health Sciences (CMHS) Institutional Review Board, University of Rwanda (Ref: CMHS/IRB/582/2024). Authorization for data collection was also obtained from the University Teaching Hospital of Butare (CHUB) Research Ethics Committee (Ref: CHUB/DG/NC/08/2098/2024). Written informed consent was obtained from all participants upon admission, authorizing the use of their medical records for research purposes. All data were handled confidentially and anonymized prior to analysis in accordance with ethical and regulatory standards.

### Study population, inclusion and exclusion criteria

The population consisted of patients hospitalized across five major departments: internal medicine (IM), surgery, gynecology (GO), pediatrics, and the intensive care unit (ICU) during the study period.

### Inclusion criteria

Department wards which admitted patients who potentially received antibiotics, being a patient who was hospitalized for disease conditions.

### Exclusion criteria

Patients who had no signed consent form in their files and patients taking long-term medication, such as tuberculosis/HIV patients, were not included in the study.

### Sample size and sampling procedure

The sample size was calculated using a single population proportion formula with a confidence interval of 95%, margin error of 3.25%, the prevalence of antibiotic prescription of 41.1% of antibiotics prescribed [19], and 10% of the non-response rate, a total of 655 patients were recruited in the study, aligning with WHO recommendation that at least 600 encounters should be included in such studies. The sample size of 655 prescriptions in all five major departments was chosen using a systematic random sampling method. Since the 4620 patients are hospitalized within one month, a sampling interval was used to recruit 655 prescriptions. The sampling interval was the number of hospitalized patients divided by the sample size (655), yielding 4620/655 = 7. As a result, every seventh patient file was chosen; however, if the selected file was from a patient who did not sign the consent form, the next file was selected.

### Data collection tool, procedure, and variables

Data were extracted from patients’ medical records at the time of patient discharge or when the patient exceeded five days of hospitalization. Data were collected using a structured data collection tool developed in alignment with the study objectives and WHO/INRUD prescribing indicators by five trained nurses who were directly involved in the dispensing of medicines within selected wards. To avoid ambiguities, the national STGs and essential medicines list were used to categorize antibiotic prescription compliance with standards [20,21].

The data collection tool comprised three sections: 1. Demographic data: age analyzed as interquartile range (IQR), gender, department of hospitalization, and medical insurance. 2. Patient history: length of admission presented in days, patient diagnosis. 3. Information related to antibiotic use, including the frequency of antibiotics prescribed, the number of antibiotics per individual prescription, the form of antibiotic administration described by number, and then antibiotics prescribed by generic names and from national essential medicine list (EML), adherence to national standard treatment guidelines (STGs) and laboratory support, availability of antibiotics was all presented as a percentage.

### Data quality control

To ensure the validity and reliability, we organized a training for data collectors before data collection, regarding the study protocol, ethical issues, and the data collection tool. A pre-test of the research tool was carried out with 5% of the sample two weeks before the actual data collection. Then, the adjustments were made based on the pre-test results, and the data collectors received refresher training. During data collection, the supervisor worked closely with data collectors to ensure data accuracy and consistency.

### Data analysis

The data was analyzed using SPSS version 25. Descriptive and analytical analysis was performed. The dependent T-test was used to analyze quantitative variables, while the chi-square was employed to evaluate the association between categorical variables. Descriptive statistics were applied to demographic information, and prescribing data were analyzed using WHO/INRUD prescribing standards, giving insight into percentage, mean, standard deviation (SD), and frequency of variables, then analytic statistics, including bivariable and multivariable regression analysis were used to identify factors associated with adherence to national STGs in antibiotic prescription. Independent variables used in regression analysis included, but were not limited to, age, gender, disease condition, use of laboratory results, department origin, and availability of guidelines. All analyses with p-values < 0.05 were considered significant.

### Results

### Demographic and clinical information

A total of 655 patients enrolled in the study through a systematic random sampling technique; the majority were female, 355 (54.2%), the average age was 35 (IQR: 10-60) years (Table 1). The highest proportion of patients were admitted to surgical wards, 211 (33%). A substantial number of patients, 562 (85.8%), had chronic infections. Additionally, prolonged hospitalization, as a stay exceeding five days, was observed among 415 (65%) patients (Table 1).

### WHO/INRUD Prescribing indicators in the prescribing survey

For 655 inpatients, antibiotics were prescribed 1265 times. The average number of antibiotics prescribed per patient was 1.95, with SD of ± 0.888 of the total antibiotics; 80% of antibiotics were prescribed from the Rwanda essential medicine list and written by generic names (Table 2). On the other hand, most of the antibiotics were written in injection form (83%). Out of 1463 inpatients in the hospital, 44.76% received at least one antibiotic, Additionally, approximately 49% of these patients were prescribed two antibiotics, with some receiving as many as four or five different antibiotics, 7% and 1% respectively (Table 2, Fig. 1). The adherence to STGs and laboratory results was 62% and 23.2%, respectively. Furthermore, the availability of antibiotics in the hospital pharmacy was 70% during the study period (see Table 2).

**Figure 1:**
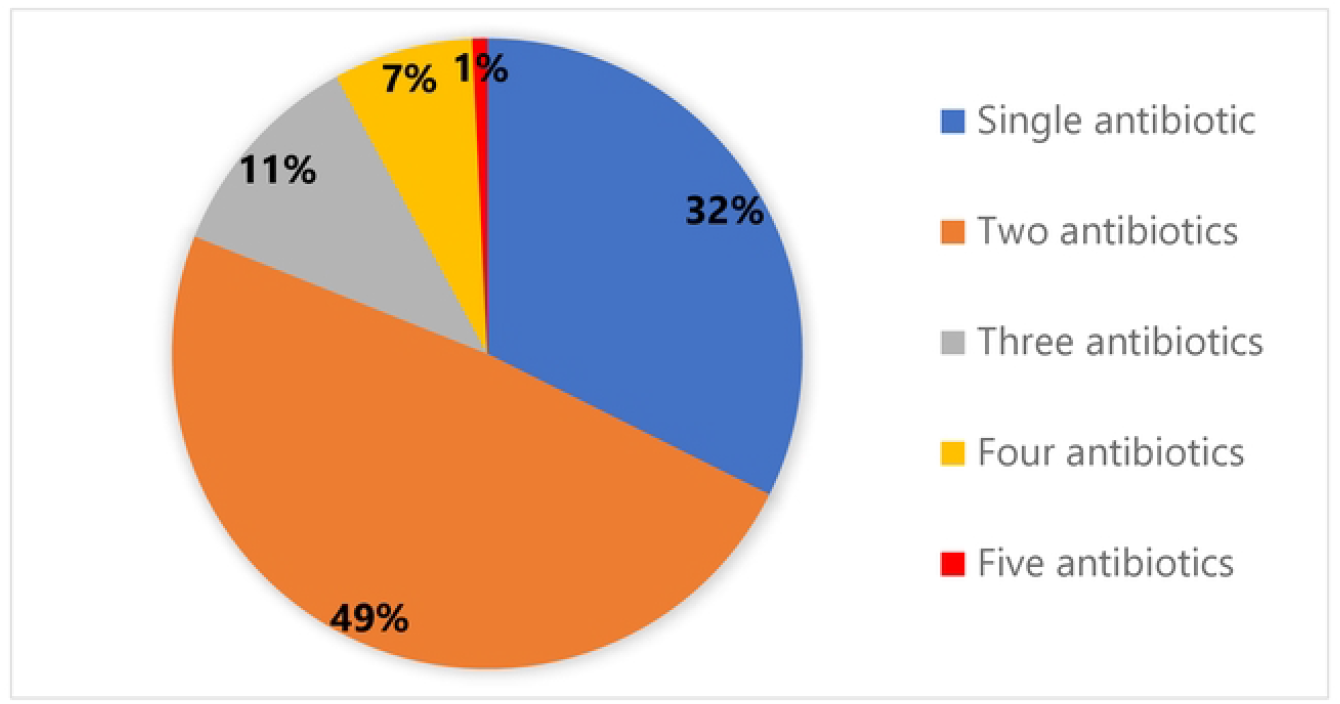
Antibiotics prescribed per patient encounter in five selected wards at the hospital in 2024.

Antibiotics that were prescribed the most were cephalosporins 520(41%) including ceftriaxone 433 (34.6%) and cefotaxime 87(6.9%); Nitroimidazoles prescribed in Flagyl name 251(20%) and penicillin (16%) (Fig. 2).

**Figure 2:**
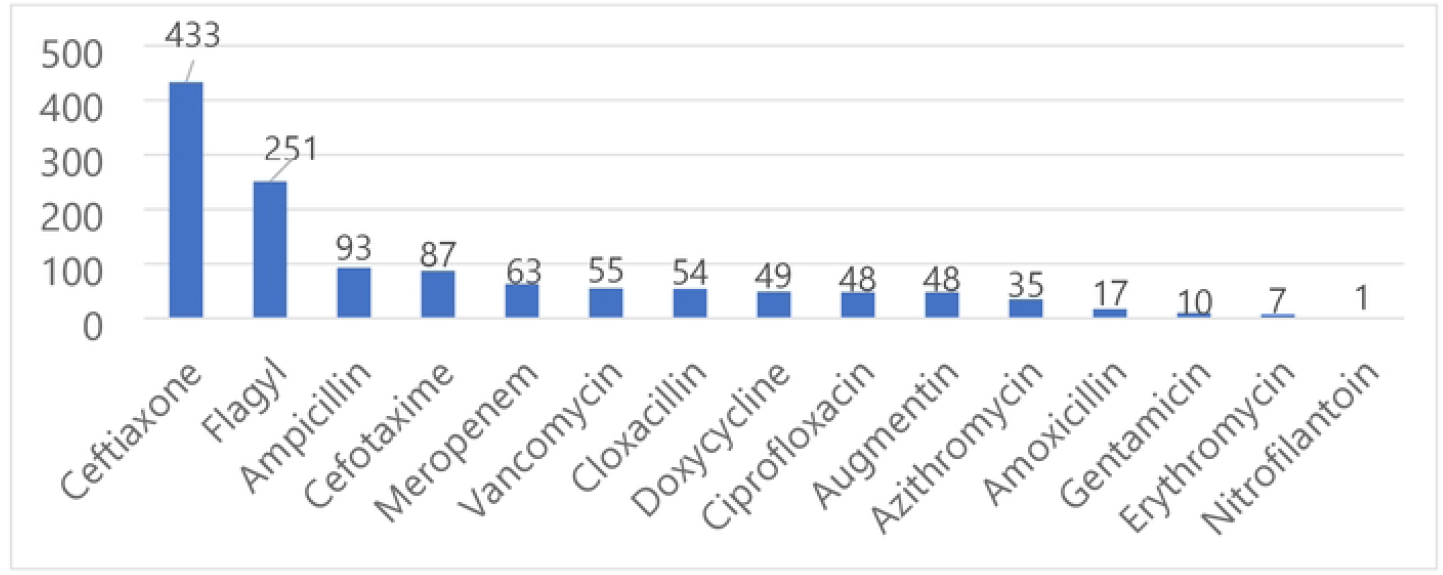
Antibiotics prescribed for admitted patients to selected wards at the hospital in 2024.

### Factors associated with irrational use of antibiotics

The analysis reveals several factors that were associated with adherence to STGs. Adherence to STGs was significantly influenced by the hospital department (Chi-square (x^2^) = 138.014, p < 0.001), patients with infections confirmed by laboratory culture and antibiogram testing (x^2^ = 64.081, p < 0.001) (Table 3). Adherence rates were substantially greater for patients with confirmed infections compared to those with unconfirmed infections. While the Patient following antibiotic therapy and adherence to STGs was found to be highly correlated (x^2^ = 28.519, p < 0.001). In addition, adherence to STGs was significantly associated with the availability of copies of the STGs and the EML (x^2^ = 38.099, p < 0.001) and (p < 0.001) (x^2^ = 53.747) respectively (Table 3).

A significant association was found between non-confirmation of infections through laboratory culture and antibiogram, and non-adherence (Adjusted OR = 0.07, p = 0.001) (Table 4). Patient adherence to the prescribed antibiotic therapy was another significant factor (Adjusted OR = 0.32, p = 0.013). Among the departments analyzed, Internal Medicine (IM) and Pediatrics (PED) demonstrated a significant deviation from STGs, with Adjusted Odds Ratios (ORs) of 0.25 (p = 0.003) and 0.33 (p = 0.002), respectively (Table 4).

## Discussion

This study aimed to assess the rational use and factors associated with antibiotic use of patients admitted to five selected departments in the hospital. To evaluate the prescription patterns, WHO/INRUD prescribing indicators were used including, the average amount of medications prescribed for each patient; the percentage of prescriptions that included an antibiotic; the percentage of prescribed antibiotics that included injections; the percentage of antibiotics prescribed under generic names, and those provided following the NEML & National STGs and an inferential analysis was performed to assess factors related to the use of antibiotics in a University Teaching Hospital.

The study showed a deviation from both WHO-endorsed standards and STGs. Additionally, most of the antibiotics were given without laboratory culture or an antibiogram. The overall average of medicines prescribed was 4.2 (SD:1.34), higher compared to the WHO prescribing standards (1.6-18), this study showed that 44.66% of patients received at least an antibiotic, indicating an overprescription, which is one of the factors of antimicrobial resistance raising globally, particularly in LMICs, calling for intervention. similar to the findings of a study conducted in Ethiopia with an average of 2.01 per prescription [22].

There was a high frequency of prescriptions for broad-spectrum antibiotics, primarily ceftriaxone and Flagyl. Similar figures were also reported in Zambia, Ethiopia, and Tanzania [19,23,24]. Conversely, a study conducted for outpatients in Tigrai, Ethiopia, reported a high use of penicillin [25]. The high frequency of broad-spectrum antibiotic prescriptions could be explained by the fact that these antibiotics were mainly prescribed without laboratory support, resulting in no evidence of infection etiology; likewise, most patients are referrals from district hospitals, commonly with chronic infections.

Furthermore, most antibiotics were administered as injections; a finding which is in line with what was observed in previous studies, including a study done in Tanzania with 78.2% being injections and in Ethiopia at 78.7% injections [19,24]. Injection rates were highest in pediatrics; this might be because children cannot easily take oral antibiotics, and injections may be considered more efficacious. However, the injections are not advised because of raised costs and increased risk of adverse drug reactions such as local infections and nerve injury [26]

This study revealed that 80% of antibiotics were written under their generic names from the national EML, with Flagyl and Augmentin being the most commonly prescribed brand names. The WHO advises that all medicines be written under generic names to reduce patient costs [10]. Similar findings were seen in studies conducted in Nigeria and Rwanda, where 82.2% and 75% of medicines were prescribed from the national EML, respectively [27,28]. On the other hand, a study conducted in Ethiopia showed a high adherence to EML, with 98.5% of antibiotics written as generic names, and 100% written from Ethiopia’s EML [24]. The low adherence to the National EML could be attributed to a lack of copies of the EML in wards and limited availability of antibiotics, as it was shown in Table2.

The study has limitations because it was only conducted in a public hospital; therefore, the findings may not apply to private health facilities. Additionally, since this study focused on inpatients where most of the antibiotics are prescribed, it might not represent outpatients. However, this research is valuable for developing strategies to promote the rational use of antibiotics and enhance the antimicrobial stewardship program in Rwanda, as it was conducted in a tertiary hospital that provides specialized services and receives largely referrals from the southern and western provinces of Rwanda.

## Conclusion

The study aimed to assess the proper use and factors linked to inappropriate use of antibiotics in tertiary hospital, we found that there is inappropriate use of antibiotics, majority of patients receive antibiotics exceeding recommended standards, and most antibiotics are written as injectable forms. There are empirical prescriptions of antibiotics and overuse of broad-spectrum antibiotics such as ceftriaxone in all studied departments. Furthermore, antibiotics prescribed without laboratory confirmation are one of the major factors of deviation from national standard treatment guidelines. Availability of antibiotics and prescription guidelines is limited in the hospital. We recommend improving adherence to diagnostic support and implementing an antimicrobial stewardship program based on evidence; however, further studies are recommended across private clinics, since this study was limited to a tertiary hospital.

## Data Availability

The data used in this study are available from the corresponding author when reasonable request and The data include patient records and prescription information collected in accordance with ethical approvals from the CMHS Institutional Review Board (Ref: CMHS/IRB/585/2024) and the University Teaching Hospital of Butare Research Ethics Committee (Ref: CHUB/DG/NC/08/2110/2024).

## Abbreviations

WHO: World Health Organization
AMR: Antimicrobial Resistance
AMS: Antimicrobial Stewardship
EML: Essential Medicine List
CHUB: Centre Hospitalier Universitaire de Butare
LMICs: Low and Middle-Income Countries
INRUD: International Network for Rational Use of Drugs
STGs: Standard Treatment Guidelines
NEML: National Essential Medicine List
ICU: Intensive Care Unit
IM: Internal Medicine
GO: Gynecology
SURG: Surgery
PED: Pediatrics
EAC RCE-VIHSCM: East African Regional Centre of Excellence for Vaccines, Immunization and Health Supply Chain Management.

## Acknowledgements

The authors express their sincere gratitude to the German Federal Ministry for Economic Cooperation and Development (BMZ) for generously funding the Master’s program in Health Supply Chain Management through the KfW Development Bank and the East African Community Regional Centre of Excellence for Vaccines, Immunization, and Health Supply Chain Management. This academic support laid the foundation for the successful execution of this research. We are also deeply thankful to the College of Medicine and Health Sciences at the University of Rwanda for their institutional support and guidance throughout the study process. Our heartfelt appreciation goes to the healthcare professionals who participated in this study—their time, insights, and dedication were invaluable. We further acknowledge the Centre Hospitalier Universitaire de Butare (CHUB) for its critical role in facilitating access for data collection.

## Authors’ contributions

OI and JBN conceived the idea. OI and JBN designed the study. OI, FH, and HS collected the data. OI did the statistical analysis. OI, JBN, KJJ, and IH wrote the manuscript. All authors read and approved the final version of the manuscript

## Ethical approval and informed consent

Ethical approval was obtained from the CMHS institutional review board (IRB), Ref: CMHS/IRB/582/2024, and Data collection approval was obtained from the University Teaching Hospital of Butare Ethics and Ethics Committee, Ref: CHUB/DG/NC/08/2098/2024. The general informed consent form was signed upon admission of patients. Therefore, this study included subjects who had signed a general informed consent form upon admission into the hospital.

## Availability of data and materials

The data used in this study are available from the corresponding author when reasonable request.

## References

1. Hutchings MI, Truman AW, Wilkinson B. Antibiotics: past, present and future. Current Opinion in Microbiology [Internet]. 2019 Oct [cited 2025 Feb 6];51:72–80. Available from: https://linkinghub.elsevier.com/retrieve/pii/S1369527419300190

2. Endale H, Mathewos M, Abdeta D. Potential Causes of Spread of Antimicrobial Resistance and Preventive Measures in One Health Perspective-A Review. IDR [Internet]. 2023 Dec [cited 2025 May 13];Volume 16:7515–45. Available from: https://www.dovepress.com/potential-causes-of-spread-of-antimicrobial-resistance-and-preventive--peer-reviewed-fulltext-article-IDR

3. WHO, WHO, AMR is a natural process that happens over time through genetic changes in pathogens. Its emergence and spread is accelerated by human activity, mainly the misuse and overuse of antimicrobials to treat, prevent or control infections in humans, animals and plants., https://www.who.int/. Antimicrobial resistance. 2023 [Internet]. 2023 mSep 21; Available from: https://www.who.int/news-room/fact-sheets/detail/antimicrobial-resistance

4. Bizimungu O, Crook P, Babane JF, Bitunguhari L. The prevalence and clinical context of antimicrobial resistance amongst medical inpatients at a referral hospital in Rwanda: a cohort study. Antimicrob Resist Infect Control [Internet]. 2024 Feb 22 [cited 2025 May 12];13(1):22. Available from: https://aricjournal.biomedcentral.com/articles/10.1186/s13756-024-01384-7

5. Salam MA, Al-Amin MY, Salam MT, Pawar JS, Akhter N, Rabaan AA, et al. Antimicrobial Resistance: A Growing Serious Threat for Global Public Health. Healthcare (Basel). 2023 Jul 5;11(13):1946.

6. Baran A, Kwiatkowska A, Potocki L. Antibiotics and Bacterial Resistance—A Short Story of an Endless Arms Race. IJMS [Internet]. 2023 Mar 17 [cited 2025 Feb 6];24(6):5777. Available from: https://www.mdpi.com/1422-0067/24/6/5777

7. Hossain MdJ, Jabin N, Ahmmed F, Sultana A, Abdur Rahman SM, Islam MdR. Irrational use of antibiotics and factors associated with antibiotic resistance: Findings from a cross-sectional study in Bangladesh. Health Science Reports [Internet]. 2023 Aug [cited 2025 Feb 6];6(8):e1465. Available from: https://onlinelibrary.wiley.com/doi/10.1002/hsr2.1465

8. Ag Ahmed MA, Ravinetto R, Diop K, Trasancos Buitrago V, Dujardin C. Evaluation of Rational Medicines Use Based on World Health Organization Core Indicators: A Cross-Sectional Study in Five Health Districts in Mauritania. IPRP [Internet]. 2024 Mar [cited 2025 Feb 6];Volume 13:17–29. Available from: https://www.dovepress.com/evaluation-of-rational-medicines-use-based-on-world-health-organizatio-peer-reviewed-fulltext-article-IPRP

9. Yimenu DK, Emam A, Elemineh E, Atalay W. Assessment of Antibiotic Prescribing Patterns at Outpatient Pharmacy Using World Health Organization Prescribing Indicators. J Prim Care Community Health [Internet]. 2019 Jan [cited 2025 Feb 6];10:2150132719886942. Available from: https://journals.sagepub.com/doi/10.1177/2150132719886942

10. WHO. Promoting rational use of medicines. 2005; Available from: https://www.who.int/activities/promoting-rational-use-of-medicines

11. Albarqouni L, Palagama S, Chai J, Sivananthajothy P, Pathirana T, Bakhit M, et al. Overuse of medications in low- and middle-income countries: a scoping review. Bull World Health Organ [Internet]. 2023 Jan 1 [cited 2025 Feb 6];101(1):36–61D. Available from: https://www.ncbi.nlm.nih.gov/pmc/articles/PMC9795388/pdf/BLT.22.288293.pdf

12. Mudenda S, Daka V, Matafwali SK. World Health Organization AWaRe framework for antibiotic stewardship: Where are we now and where do we need to go? An expert viewpoint. ASHE [Internet]. 2023 [cited 2025 Feb 6];3(1):e84. Available from: https://www.cambridge.org/core/product/identifier/S2732494X2300164X/type/journal_article

13. Bassetti S, Tschudin-Sutter S, Egli A, Osthoff M. Optimizing antibiotic therapies to reduce the risk of bacterial resistance. European Journal of Internal Medicine [Internet]. 2022 May [cited 2025 May 13];99:7–12. Available from: https://linkinghub.elsevier.com/retrieve/pii/S0953620522000395

14. Aljofan M, Oshibayeva A, Moldaliyev I, Saruarov Y, Maulenkul T, Gaipov A. The rate of medication nonadherence and influencing factors: A systematic Review. ELECTRON J GEN MED [Internet]. 2023 May 1 [cited 2025 May 13];20(3):em471. Available from: https://www.ejgm.co.uk/article/the-rate-of-medication-nonadherence-and-influencing-factors-a-systematic-review-12946

15. Siahaan S, Rukmini R, Roosihermiatie B, Andarwati P, Handayani RS, Tarigan IU, et al. The Effort to Rationalize Antibiotic Use in Indonesian Hospitals: Practice and Its Implication. Khamesipour F, editor. Journal of Tropical Medicine [Internet]. 2023 Feb 25 [cited 2025 Feb 6];2023:1–12. Available from: https://www.hindawi.com/journals/jtm/2023/7701712/

16. Igizeneza A, Bitunguhari L, Masaisa F, Hahirwa I, Uwamahoro LD, Sebatunzi O, et al. Prescription Practices and Usage of Antimicrobials in a Tertiary Teaching Hospital in Rwanda: A Call for Antimicrobial Stewardship. Antibiotics [Internet]. 2024 Oct 31 [cited 2025 Feb 14];13(11):1032. Available from: https://www.mdpi.com/2079-6382/13/11/1032

17. Nkurunziza Sebuhoro Dieudonne3, Tihon Vincent4, Muganga, Raymond. Appropriateness of Antibiotic Prescription Practices in Health Centers in the District of Gisagara, Rwanda. 2022;Rw. Public Health Bul. Vol. 3 (2); December 2022.(December 2022.). Available from: https://rbc.gov.rw/publichealthbulletin/img/rphb_issues/247ce19aa59c37296375beaa99fcc3051673256214.pdf

18. Munyemana JB, Gatare B, Kabanyana P, Ivang A, Mbarushimana D, Itangishaka I, et al. Antimicrobial Resistance Profile of Bacteria Causing Pediatric Infections at the University Teaching Hospital in Rwanda. The American Journal of Tropical Medicine and Hygiene [Internet]. 2022 Dec 14 [cited 2025 May 13];107(6):1308–14. Available from: https://www.ajtmh.org/view/journals/tpmd/107/6/article-p1308.xml

19. Costantine JK, Bwire GM, Myemba DT, Sambayi G, Njiro BJ, Kilipamwambu A, et al. WHO/INRUD prescribing indicators among tertiary regional referral hospitals in Dar es Salaam, Tanzania: a call to strengthen antibiotic stewardship programmes. JAC-Antimicrobial Resistance [Internet]. 2023 Jul 11 [cited 2025 Feb 6];5(4):dlad093. Available from: https://academic.oup.com/jacamr/article/doi/10.1093/jacamr/dlad093/7236566

20. MOH. RWANDA STANDARD TREATMENT GUIDELINES [Internet]. 2022. Available from: https://www.moh.gov.rw/index.php?eID=dumpFile&t=f&f=92525&token=65bb1585f37424835c9eeb4c9ad4747b38a80514

21. WHO. Rwanda: National List of Essential Medicines for Adults 2022 (English) [Internet]. Vol. 7th Edition, Volume 14. 2024. Available from: https://www.who.int/publications/m/item/rwanda--national-list-of-essential-medicines-for-adults-2022-(english)

22. Demoz GT, Kasahun GG, Hagazy K, Woldu G, Wahdey S, Tadesse DB, et al. Prescribing Pattern of Antibiotics Using WHO Prescribing Indicators Among Inpatients in Ethiopia: A Need for Antibiotic Stewardship Program. Infect Drug Resist. 2020;13:2783–94.

23. Chizimu JY, Mudenda S, Yamba K, Lukwesa C, Chanda R, Nakazwe R, et al. Antibiotic use and adherence to the WHO AWaRe guidelines across 16 hospitals in Zambia: a point prevalence survey. JAC-Antimicrobial Resistance [Internet]. 2024 Sep 3 [cited 2025 Feb 24];6(5):dlae170. Available from: https://academic.oup.com/jacamr/article/doi/10.1093/jacamr/dlae170/7841613

24. Tadesse TY, Molla M, Yimer YS, Tarekegn BS, Kefale B. Evaluation of antibiotic prescribing patterns among inpatients using World Health Organization indicators: A cross-sectional study. SAGE Open Medicine [Internet]. 2022 Jan [cited 2025 Feb 6];10:20503121221096608. Available from: https://journals.sagepub.com/doi/10.1177/20503121221096608

25. Hailesilase GG, Welegebrial BG, Weres MG, Gebrewahd SA. WHO/INRUD prescribing indicators with a focus on antibiotics utilization patterns at outpatient department of Adigrat general hospital, Tigrai, Ethiopia: a retrospective cross-sectional study. Antimicrob Resist Infect Control [Internet]. 2024 Nov 6 [cited 2025 May 16];13(1):133. Available from: https://aricjournal.biomedcentral.com/articles/10.1186/s13756-024-01490-6

26. Chang Y, Chusri S, Sangthong R, McNeil E, Hu J, Du W, et al. Clinical pattern of antibiotic overuse and misuse in primary healthcare hospitals in the southwest of China. Angelillo IF, editor. PLoS ONE [Internet]. 2019 Jun 26 [cited 2025 Feb 6];14(6):e0214779. Available from: https://dx.plos.org/10.1371/journal.pone.0214779

27. Ntirenganya C, Manzi O, Muvunyi CM, Ogbuagu O. High Prevalence of Antimicrobial Resistance Among Common Bacterial Isolates in a Tertiary Healthcare Facility in Rwanda. The American Society of Tropical Medicine and Hygiene [Internet]. 2015 Apr 1 [cited 2025 Feb 6];92(4):865–70. Available from: https://www.ajtmh.org/view/journals/tpmd/92/4/article-p865.xml

28. Oli AN, Onyeaso N, Emencheta SC, Ofomata CM, Kretchy JP, Okhamafe A, et al. Evaluating antimicrobial prescribing in a Tertiary Healthcare Institution in Nigeria. J of Pharm Policy and Pract [Internet]. 2021 Dec [cited 2025 Feb 6];14(1):99. Available from: https://joppp.biomedcentral.com/articles/10.1186/s40545-021-00380-1

